# When thinking you are better leads to feeling worse: Self-other asymmetries in pro-social behavior and increased anxiety during Covid-19

**DOI:** 10.1101/2021.02.26.21252547

**Authors:** Chelsea Helion, Virginia Ulichney, David V. Smith, Johanna Jarcho

## Abstract

Self-favoring beliefs (e.g., that one tends to perform better than peers) are generally associated with positive psychological outcomes like increased self-esteem and resilience. However, this tendency may be problematic in the context of collective action problems, wherein individuals are reliant on others’ pro-social behaviors to achieve larger goals. We examined this question in the context of the Covid-19 pandemic, and recruited participants (*n* = 1023) from a university community in Spring 2020. We found evidence for a self-peer asymmetry, such that participants reported that they were doing more to stop the spread of the disease and were more pro-socially motivated than peers. Actual peer reports indicated that these were overestimations. This self-enhancement tendency comes with a cost: the perceived self-peer asymmetry mediated the relationship between Covid-specific worry and general anxiety during the early lockdown period. This indicates that while believing one is doing more than others may be maladaptive in collective action problems.

## Introduction

Whether it’s thinking we are better drivers, better partners, or simply better people, we tend to evaluate ourselves more favorably than we do others (Kruger, 1999; Ross & Sicoly, 1979; Epley & Dunning, 2000). This bias potentially creates a psychological tension in densely networked societies, wherein we often *have* to rely on others to progress in any domain (Fitzsimons, Finkel, & vanDellen, 2018). What’s more, people tend to self-enhance on the values that they prioritize, and these personally-held values can in turn be influenced by broader societal principles. For example, Sedikides, Gaertner, & Toguchi (2003) found that, when engaging in social comparison, those in the United States (where independence tends to be more societally prioritized) *and* anyone who prioritized independence often played up their own individualist traits relative to others. In contrast, people in Japan (where interdependence tends to be more societally prioritized) *and* individuals who valued interdependence alike tended to inflate their personal collective-focused attributes compared to others (Fatehi, Priestley, & Taasoobshirazi, 2020). Especially in environments wherein individualism is societally prioritized (such as the U.S.), if people are surrounded by those they tend to perceive as less competent, less knowledgeable, and more selfish than themselves (Pronin, Lin, & Ross, 2002), they may also be pessimistic about the likelihood of collective action to bring about positive change. For example, in the United States uniquely, people tend to find their own violations of protective Covid-19 protocols (e.g., social distancing, mask-wearing) more acceptable than others’, a perception which is not shared, for example, by their counterparts in China (Dong et al., 2021).

The Covid-19 epidemic reflects this tension more than any other time in recent memory — seldom have we been so reliant on others’ behaviors to quickly produce positive societal outcomes. As such, those who fail to act on behalf of collective protection (e.g., failure to vaccinate, provided adequate access to do so) consequently tend to incur reduced generosity from those who choose to act in the interest of the collective (e.g., by vaccinating; Weisel, 2021). Although action to mitigate the impact of a societal challenge may be enhanced by some social motivations (e.g., norms, moralization), it also may be diminished through social comparison as self-serving biases cause one to inaccurately believe they are doing more to mitigate the issue than peers (Ajzen, 1991). While self-serving biases can be adaptive, they may be psychologically suboptimal in situations that require relying on others for positive outcomes, i.e., collective action problems. We test this hypothesis by examining whether, in the setting of a U.S. university community, the individuals who are most likely to think they do better than their peers are also the ones that psychologically fare worse when they are in a situation (i.e., the Covid-19 pandemic) wherein achieving a positive outcome requires reliance on the behaviors of others.

Self-serving bias is associated with higher self-esteem (Campbell, Rudich, & Sedikides, 2002), underlies predictions for rosier futures (Kruger & Burrus, 2004), and dulls recollections of less-than-ideal pasts (Steimer, Mata, & Simao, 2009). However, this tendency can also lead to undue cynicism (Kruger & Gilovich, 1999) and result in inaccurate perceptions of both ourselves and others (Sedikides, Campbell, Reeder, & Elliott, 1998). For example, individuals claim more responsibility for joint outcomes than is logically possible (Ross & Sicoly, 1979; Kruger & Gilovich, 1999). This kind of self-enhancement is generally benign -- absent the occasional argument with a partner that might come from overclaiming one’s contribution to the housework, these biases may typically boost positivity and increase productivity (Taylor & Brown, 1988). However, they may become problematic when positive societal outcomes depend on the behaviors of others. If individuals are overly confident in their ability to predict others’ behavior (Vallone, Griffin, Lin, & Ross, 1990; Pronin, 2009), but those predictions are overly cynical (Kruger & Gilovich, 1999), then outcomes that rely on others doing the “right thing” may seem unlikely to occur. Individuals who think more of themselves and less of their peers may thus be particularly pessimistic about the likelihood of resolving collective action issues.

The Covid-19 pandemic presents an important example of a collective action problem. To successfully contain the virus, individuals have to collectively practice novel behaviors like mask-wearing and social-distancing (CDC, 2020). Some have posited that one of the most powerful predictors of collective action success in Covid-19 is trust (Van Bavel et al., 2020) -- both horizontal trust in each other (Lenton, Boulton, & Scheffer, 2022) and vertical trust in our institutions (Harring, Jagers, & Löfgren, 2020). The former -- the trust that we place in others to do the right thing for the benefit of the group -- is the focus of the research presented below. We hypothesize that individuals will report increased engagement in behaviors that reduce the spread of Covid-19 relative to their peers. Moreover, we predict that this lack of trust in peers will be related to increased Covid-related concern (e.g., stress, worry), and be associated with heightened anxiety during the pandemic. It is important to note that stress and worry are conceptualized in this article as fleeting experiences that arise in distinct situations and are not necessarily indicative of anxiety on their own (Zebb & Beck, 1998). Anxiety is considered a chronic, enduring experience characterized by chronically excessive levels of these negative affective experiences that can result in a clinical diagnosis at its most severe (Bystritsky et al., 2013; Borkovec & Inz, 1990).

We examine two areas of predicted self-other asymmetry with regard to the Covid-19 pandemic: 1) that individuals will report being more knowledgeable about Covid-19 than their peers (Hypothesis 1), and 2) that individuals will report having made more Covid-19 behavioral changes relative to their peers (Hypothesis 2). Turning first to Covid-related knowledge, individuals tend to overestimate their knowledge in domains where absolute knowledge is high relative to domains where it is low (Kruger et al., 2008). As the pandemic had received a great deal of coverage in the media at the time of data collection (Pew Research Center, 2020), we predicted that Covid-19 may be a domain where absolute knowledge is relatively high, and that individuals would subsequently overestimate their knowledge relative to peers.

Second, Covid-19 related behaviors are morally-relevant. At the time of data collection, individuals who were more pro-socially inclined report wearing a mask more frequently and perceived others who wore masks as being more pro-social than those who did not (Betsch et al., 2020). This suggests that Covid-19-related behavior is moralized, and would have meaningful implications for self-other asymmetry predictions, as individuals tend to make highly self-serving asymmetric predictions about actions that have moral implications (Epley & Dunning, 2000). We predicted that individuals would report restricting their behavior to a greater extent than peers, and moreover, would be more likely to endorse doing so for pro-social as compared to self-focused reasons.

We propose lastly that the extent to which individuals think their peers are ill-informed and are doing less to protect others than they are is associated with increased anxiety during the lockdown period (Hypothesis 3). Anxiety symptoms have been highly prevalent during the Covid-19 pandemic (Salari et al., 2020; Huang & Zhao, 2020; Ahmed et al., 2020; Bäuerle et al., 2020). There are many possible reasons for this increase, including increased media and news consumption (Moghanibashi-Mansourieh, 2020), economic concerns (Mann, Krueger, & Vohs, 2020), and concern about individual/familial health (World Health Organization, 2020). We posit that another contributor may be the lack of control individuals have over the behaviors of others paired with the centrality of others’ behaviors in determining Covid-19 related outcomes. We hypothesized that individuals who show a larger self-other asymmetry in Covid-related knowledge and behavior would report higher levels of both Covid-specific worry, general anxiety symptoms, and stress during the initial lockdown period.

Notably, prior research has found a *negative* relationship between the tendency to self-enhance relative to one’s peers and anxiety (Hoorens & Buunk, 1993). However, we predict the opposite effect in this context, given that surviving a global pandemic necessitates reliance on others. If individuals believe that their peers are doing less than they are to contain the spread of Covid-19, then they may feel more worried and stressed about Covid-19, and thus more anxious during the lockdown period. To examine this question, we tested two indirect effects models wherein the size of self-peer asymmetry in Covid-19-related restrictive behaviors mediated the relationship between 1) Covid-specific worry and 2) stress with general anxiety reported during the early stages of the Covid-19 pandemic.

## Methods

The data presented here is part of the first wave of a longitudinal study collected at a large northeastern university in the U.S with Institutional Review Board approval. The first wave of data collection was completed on 3/25/20-4/4/20, approximately 2-3 weeks following the closure of the university on 03/11/2020, and consisted of 1852 participants (1170 Female, *M*_*age*_ = 24.03, *SD*_*age*_ *=* 9.24, age range = 18 - 76). Given the long-term longitudinal aims, we recruited as many participants as possible for the first wave. The total sample consisted of 1386 undergraduate students (74.84%), 166 graduate students (68 Masters students, 98 PhD students; 8.96%), 142 staff members (7.67%), 80 faculty members (43 Non-Tenure Track, 16 Tenure-Track; 4.32%), 26 individuals who identified as “Other” (1.4%), and 52 (2.81%) who did not indicate the nature of their affiliation with the university. We only included participants who filled out (rather than leaving unanswered or selecting “prefer not to respond”) all of the measures of interest (Covid-related behavioral restriction and risk-assessment), leaving us with a final sample of 1023 participants (697 females, *M*_*age*_ = 23.61, *SD*_*age*_ = 8.52, age range = 18 - 71). We believe that this sample size is appropriate given that all analyses were conducted within-participant. Undergraduate participants were compensated through course credit based on department-level acceptance of credits. All participants were entered into a raffle at each wave of the study. Participants were recruited via university listservs, and their participation entered them in a lottery to win a $100 Amazon gift card.

All participants were members of the larger university community (e.g., undergraduates, graduate students, faculty, etc.). This is perhaps an ideal sample in which to study the nature and accuracy of self-peer asymmetries as we can compare individual estimates of the average population with what the average population is actually reporting. For example, we can compare what an individual undergraduate believes their peers are doing with what undergraduate students are actually reporting in the sample. This provides an ecologically-valid, concrete point of reference from which to examine whether individuals’ beliefs about where they stand relative to their peers are accurate.

### Measures

We collected a number of measures on this sample, but for the purposes of this manuscript, only focused on the variables listed here. When asking participants to recall behaviors, we asked specifically about the past two weeks as that approximately matched up to the initiation of Covid-19 protocols at the university level and ensured consistency across both participants’ considerations and measures (e.g., validated clinical anxiety and stress scales ask about the past two weeks). Unless indicated otherwise (e.g. explicitly exploratory analyses), these analyses were preregistered on AsPredicted.org: https://aspredicted.org/blind.php?x=/HGQ_8XB.

#### Covid-19 Knowledge

Accuracy of Covid-19 symptoms and general knowledge were evaluated based on the information that was known at the time -- March/April 2020. We acknowledge that knowledge about common symptomology for Covid-19 has changed as medical researchers learn more about the disease (Burke *et al*., 2020).

### Perceived self-peer Covid-19 knowledge asymmetry

To assess perceived relative knowledge regarding Covid-19, participants were asked to indicate “how knowledgeable they were about Covid-19 compared to their peers” on a scale from 0 = “Know much less” to 6 = “Know much more”. On this scale, the midpoint (3) would indicate an assessment of equivalent knowledge relative to peers. This measure asked directly whether participants felt that they knew more about Covid-19 than peers. As such, we could uniquely use this measure as an indicator of perceived self-peer Covid-19 knowledge asymmetry rather than calculating a difference score between separately-reported self and peer estimates as we do for other measures (e.g., Covid self-peer risk assessment).

### Actual Covid-19 symptom knowledge

To assess actual knowledge relative to one’s peers, we asked participants to indicate the three most common symptoms of Covid-19 from the following list (the correct answers – symptoms most commonly associated with the wave of Covid-19 infecting the population in March/April 2020 - have been bolded): fatigue, nausea, **shortness of breath**, sneezing, diarrhea, constipation, **cough**, runny nose, **fever**, body aches, and ulcers in the mouth; Burke et al., 2020). For each participant, we summed the number of symptoms that they correctly identified and the number of symptoms that they incorrectly identified, and created a difference score by subtracting the incorrect number from the correct number, such that higher numbers indicate greater knowledge of Covid-19 symptomatology.

### Covid-19 general knowledge

To further assess Covid-related knowledge at a time in which it was still novel and wherein it could be challenging for the public to delineate between accurate and inaccurate information (Van Bavel et al., 2020), participants were asked to read each of the following statements, and to indicate the extent to which they believed these statements about Covid-19 (correct statements as identified at the time of data collection have been bolded): 1) **“Someone can transmit it, even if they do not show symptoms”**, 2) “Your dog or cat can transmit it to you”, 3) **“Standard surgical masks prevent you from becoming infected”**, 4) “It mutated from the common cold”, 5) “It was probably made in the lab”, 6) “It is less deadly than the annual flu”, 7) “You can become infected from eating at a Chinese restaurant”, 8) “Vitamin C supplements will help keep you from being infected”. All statements were evaluated on a scale from 0 = “Proven False” to 6 = “Proven True”. For each participant, we calculated their average response for the correct items and their average response for the incorrect items, and created a difference score by subtracting their incorrect average from their correct average, such that higher numbers indicate greater knowledge of Covid-19.

### Assessing self-peer knowledge asymmetries

To examine whether an assessment of being more or less knowledgeable about Covid-19 reflected reality, we calculated the average Covid-19 symptom knowledge and average Covid-19 transmission knowledge for each peer group in the sample (i.e., undergraduates, graduate students, faculty, etc.). We then compared the individual participant’s averages to their peer group averages. To do so, we subtracted the peer average from each participant’s knowledge score, such that higher numbers indicate *actually* being more knowledgeable about Covid-19 relative to one’s peers. The difference score for symptom-knowledge was moderately skewed (skewness = - .97), so to examine whether a predicted asymmetry in Covid-19 related knowledge was associated with actual symptom knowledge, we calculated Spearman’s rank correlation between these two variables.

#### Covid-19 Behavior

##### Risk Assessment

###### Covid self-peer risk assessment

Participants were asked to estimate their risk of becoming infected with Covid-19, their risk of transmitting Covid-19, and their risk of dying if they were to become infected with Covid-19. They were then asked to make the same assessments for their average peer. All risk estimates were between 0-100%. To examine how perceived self-focused (infection risk) vs. social-focused (transmission risk) COVID-risk assessment differed across the self and other, we ran an exploratory repeated-measures 2 (target: self, other) x 2 (action-type: infection, transmission) ANOVA using the “rstatix” package (Kassambara, 2020) in R.

##### Behavioral Restriction

###### Self- and peer reported behavioral restriction

Participants were asked to indicate how much they had limited their social interactions to reduce their likelihood of becoming infected (self-focused) and how much they had limited their social interactions to reduce their likelihood of exposing others to Covid-19 (pro-social-focused). They were also asked to indicate to what extent they felt that their average peer had done the same. All questions were answered on a scale from 0 = “Not at all” to 6 = “Extremely”.

###### Specific behavior change

To assess specific behavior change, we asked participants to indicate the number of behaviors that were common pre-pandemic (e.g., going to restaurants, attending events with audiences, using ride share services) that they had limited within the past two weeks from a list of 21 behaviors (see Supplemental materials for full list of behaviors). We created a count variable for the number of behaviors that the participants had reported limiting, such that higher numbers indicate limiting more behaviors.

#### Worry, Stress, & Anxiety Scales

##### Covid-related worry

Participants were asked to indicate how often they worried about Covid-19 during the past two weeks via the following questions (all on a scale from 0 = “Not at all” to 6 = “Extremely frequently”): “I worried about my health”, “I worried about Covid-19”, “I worried about all the things I could do about Covid-19”, “News about all health-related topics made me worry about Covid-19”, “Changing my routine made me worry about Covid-19”, “I talked with my friends about Covid-19”, and “I felt like I had control over whether or not I would become infected with Covid-19 (reverse-scored).” These questions had good internal consistency, *a* = .81, and were combined to create a composite variable of Covid-19 related worry.

##### Stress

Stress was assessed using the Perceived Stress Scale (PSS; Cohen, Kamarck, & Mermelstein, 1983). Participants indicated how often they had experienced stress over the past two weeks, with higher numbers on the scale reflecting more stress.

##### Anxiety

Symptoms of anxiety were assessed using the anxiety subscale of the DSM-5 Cross-Cutting Symptom Measure (DSM-5-CC; APA, 2013). Higher numbers reflect more severe anxiety over the past two-week period.

### Analysis

#### Quantifying self-peer asymmetries

##### Relationship between predicted behavioral restriction and actual behavioral restriction

To examine whether the predicted asymmetry in predicted behavioral restriction (via difference score between self and peer) was associated with an asymmetry in actual behavioral restriction (via difference score between self and peer), we conducted an exploratory Spearman’s rank correlation between predicted behavioral restriction and actual behavioral restriction. We selected this method as the actual behavior restriction variable was highly skewed (skewness = - 1.58). For both predicted and actual behavioral restriction variables, higher scores indicated greater personal than peer restriction, and lower scores indicated greater peer than personal restriction. As such, a stronger positive correlative relationship would indicate that people both expected, and truly did take more action than peers as predicted. Likewise, a negative correlative relationship would indicate that expectations of greater personal than peer action were not consistent with reality, and instead, individuals with high personal expectations actually took fewer actions than peers (and vice-versa).

##### Relationship between perceived behavioral restriction and specific behavior change

To examine whether perceived differences in behavioral restrictions relative to one’s peers translates to specific behavior change, we ran an exploratory linear regression with predicted behavioral restriction as the predictor variable, and specific behavior change as the outcome variable.

##### Relationship between self-peer asymmetries and mental health outcomes

To examine the role that a perceived self/other asymmetry may play in maladaptive emotional responding we conducted the following analyses.

#### Knowledge asymmetries and mental health outcomes

We examined the relationship between our asymmetry measures (knowledge asymmetry, behavioral restriction asymmetry) and both Covid-related worry and general anxiety. We first ran a preregistered simple linear regression with Covid-related knowledge asymmetry predicting Covid-related worry. A positive relationship would indicate that individuals who believe that they know more about Covid-19 relative to their peers are also more worried about Covid-19. To examine whether knowledge asymmetry predicts Covid-19 worry specifically or stress and anxiety more generally, we also ran two preregistered linear regressions with covid-related knowledge asymmetry predicting stress (as assessed by the PSS) and anxiety (as assessed by the DSM-CC-5).

#### Behavioral restriction asymmetries and mental health outcomes

To examine whether believing one is restricting behavior more than peers is related to negative mental health outcomes, we ran a preregistered simple linear regression with the predicted behavioral restriction as a fixed factor predicting Covid-specific worry. To examine whether asymmetries in perceived behavioral restriction predicts Covid-specific worry specifically or anxiety more generally, we also ran two preregistered linear regressions with predicted behavioral restriction predicting perceived stress and anxiety, respectively, over the past two weeks.

#### Testing whether perceived self-peer asymmetry mediates the relationship between Covid-specific worry and general anxiety

Next, to examine whether a larger self-other asymmetry is linked to more negative mental health outcomes during the Covid-19 lockdown period, we conducted an exploratory mediation analysis to test whether the degree of predicted self-other behavioral restriction mediated the relationship between Covid-specific worry and general anxiety during the lockdown period. We used the averaged behavioral restriction asymmetry score (i.e., the average of the predicted self-other self- and pro-social-focused behavioral restriction estimates). Using the “lavaan” package in R (Rosseel, 2012) we estimated two indirect effects models, testing whether the size of the predicted self-other asymmetry mediated the relationship between 1) Covid-specific worry and 2) stress and reported general anxiety (respectively).

## Results

### Self-other asymmetries in Covid-19 knowledge (Hypothesis 1)

#### Individuals are overconfident about Covid-19 knowledge relative to peers

We first hypothesized that individuals would report knowing more than their average peer about Covid-19, but that this would not necessarily track with reality. A one-sample t-test on perceived Covid-19 knowledge relative to one’s peers (i.e., perceived Covid-19 knowledge asymmetry) found that the mean (*M* = 3.48, *SD* = 1.15) was significantly different from the midpoint (3), *t*(1022) = 13.48, *p* < .001, *d* = .42, indicating that our sample believed that they knew more than their peers about Covid-19. A paired t-test indicated that participants were more likely to endorse correct transmission knowledge about Covid-19 (*M* = 2.56, *SD* = .59) as compared to incorrect symptom knowledge (*M* = .52, *SD* = .73), *t*(1022) = 54.31, *p* < .001, *d* = 1.7. However, perceived Covid-19 knowledge asymmetry was not related to being more knowledgeable about Covid-19 symptoms: *r*_*s*_ = .044, 95% CI: [-.017, .105], *p* = .159 (examined via Spearman’s rho due to violation of the normality assumption, skewness _SymptomDifference_ = -0.97, necessary for the planned linear regression), or general Covid-19 knowledge: *b* = .015, *se* = .030, 95% CI: [-.045, .075], *t*(1020) = .49, *p* = .625, relative to one’s peers. Taken together, this indicates that while individuals believed that they are more knowledgeable about Covid-19 than their peers, this belief did not reflect actually knowing more about Covid-19 symptoms or how the disease spreads.

#### Individuals are optimistic about their likelihood of infection relative to peers

We hypothesized that, consistent with other research on unrealistic optimism regarding health-related risk (Weinstein, 1982), participants would indicate that their peers were at higher risk than they were for getting infected with (i.e., self-focused) and transmitting Covid-19 (i.e., pro-social) to others. We found a significant target (2: self, peer) by action-type (2: infection, transmission) interaction, *F*(1, 4084) = 11.35, *p* < .001, partial eta-squared = .003. Post-hoc pairwise comparisons of this interaction indicated that in the self condition, the perceived likelihood of transmitting the virus to others was higher (*M* = 36.88, *SD* = 30.95) than perceived likelihood of getting infected (*M* = 32.63, *SD* = 24.31), *t*(1021) = 5.29, *p*_*adj*_ < .001. This was smaller than the difference observed in the peer condition, wherein the predicted likelihood of transmitting the virus to others was higher (*M* = 52.2, *SD* = 29.81) than the perceived likelihood of getting infected (*M* = 42.09, *SD* = 25.53), *t*(1021) = 15.26, *p*_*adj*_ < .001) (Figure 1). We found no differences in estimated risk of death following COVID-19 infection between self (*M* = 11.28, *SD* = 17.53) and peer (*M* = 11.41, *SD* = 15.95), *t*(1022) = .29, *p* = .77 *d* = 0.009, suggesting that our participants believed that they were as likely as peers to be negatively impacted by Covid-19, were they to contract it.

**Figure 1.**
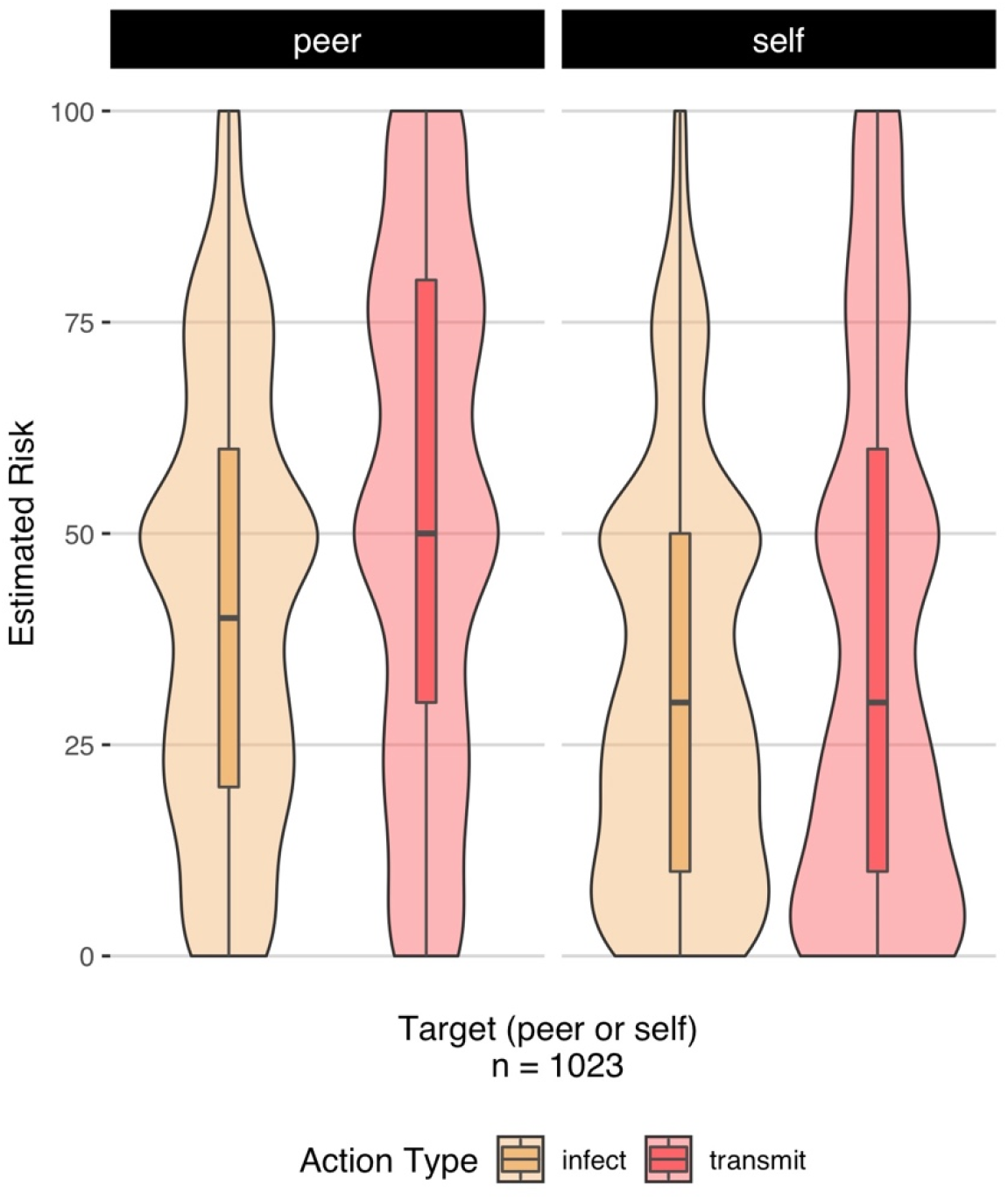
Interaction between Target (self, other) and Action Type (Getting infected with Covid-19, Transmitting Covid-19 to someone else) on estimated risk. Relative to their peers, individuals predicted that they would be less likely to get infected with Covid-19, and also less likely to transmit it to others.

### Self-other asymmetries in perceived Covid-19 behavior (Hypothesis 2)

#### Individuals believe that they are restricting their behavior more, and for more pro-social reasons, relative to peers

Turning first to beliefs about behavior change, we found a significant interaction between target (self, peer) and action type (infection, transmission) on perceived behavior change, *F*(1,4088) = 5.19, *p* = .023, partial eta-squared = .001. Post-hoc pairwise comparisons of this interaction indicated that individuals were more likely to report that they had limited their behaviors to avoid transmitting the virus to others (*M* = 5.24, *SD* = 1.21), as compared to protecting themselves from infection (*M* = 5.14, *SD* = 1.18), *t*(1023) = 2.92, *p* = .004. In contrast, they reported the *opposite* for their peers, such that they estimated that their peers were less motivated to limit their behaviors to avoid transmitting Covid-19 to others (*M* = 3.40, *SD* = 1.42), as compared to protecting themselves from infection (*M* = 3.48, *SD* =1.25), *t*(1023) = 3.07, *p* = .002 (Figure 2). Taken together, this indicates that individuals believe that they limited their behavior in order to protect others, but that their peers had done so to protect themselves.

**Figure 2.**
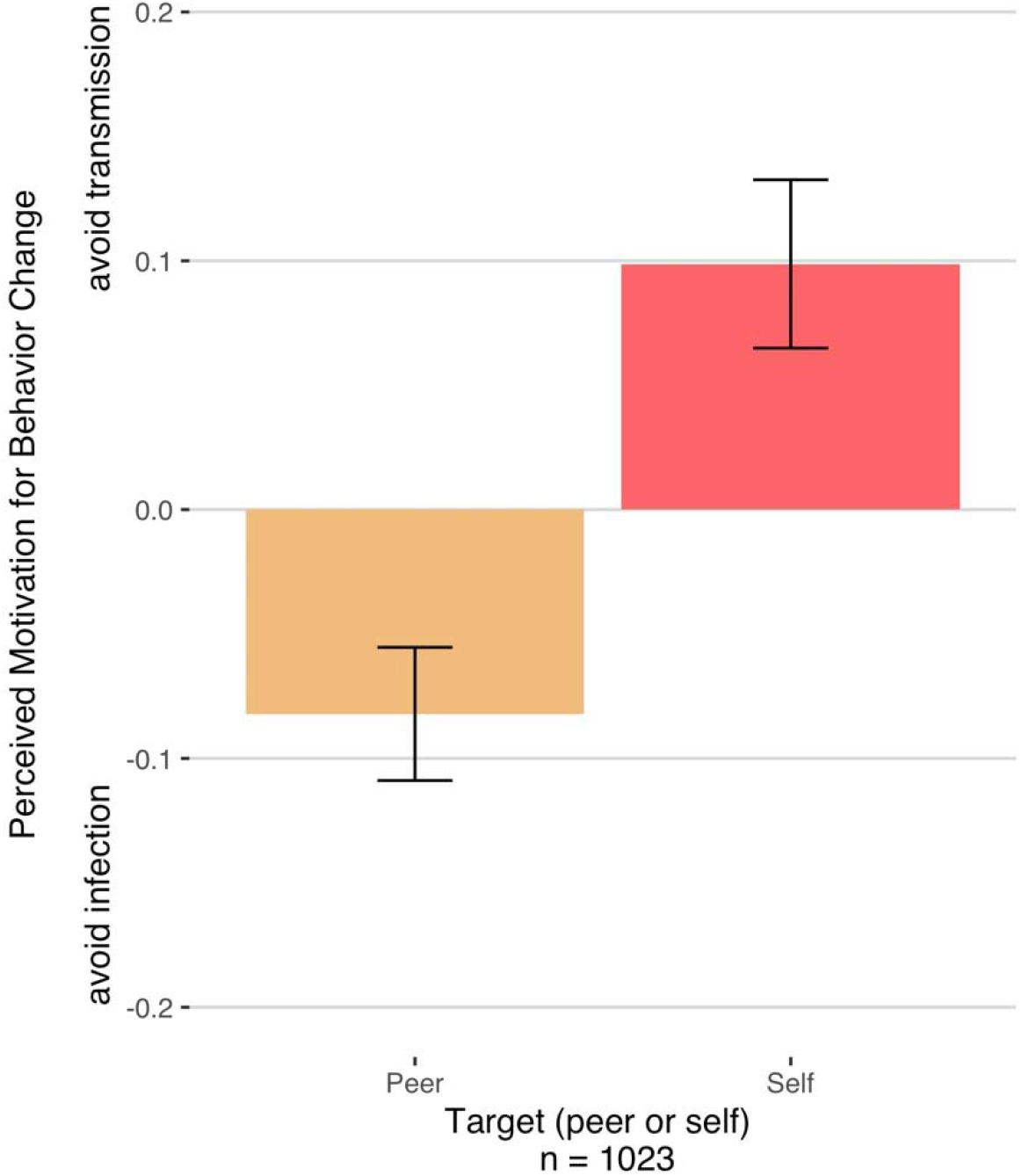
Interaction between Target (Self, Peer) and Perceived Motivation for Behavior Change (restricting behavior to avoid infection vs. restricting behavior to avoid transmission).

Individuals reported that they had limited their behaviors to avoid transmitting the others more than they had limited it to avoid getting infected. They reported that the opposite had been the case for their peers.

#### Are beliefs about behavioral restriction relative to peers accurate?

To examine whether these assessments tracked with reality, we computed 3 difference scores, outlined in Table 1. Given that they were highly correlated (*r* = .76), we collapsed across transmission-based and infection-based restriction for the remaining analyses. For all scores, higher numbers reflect a bias towards the self restricting more than peers (Table 1).

**Table 1.** Difference score calculations to assess each of the measured self-peer asymmetries.

| Asymmetry Type | Variables |
| --- | --- |
| Predicted Behavioral Restriction | (Self-reported behavioral restriction -- self-estimated peer restriction) |
| Actual Behavior Restriction | (Self-reported behavioral restriction – average peer restriction) |
| Specific Behavior Change | (Number of self-reported specific behaviors -- Number of average peer specific behaviors) |

#### Individuals are inaccurate about magnitude, but right about direction

Due to the nature of our sample, we can again test whether individuals’ beliefs about their average peer hit the mark. Given that the process through which the reported self-peer discrepancy was calculated (see Methods) was largely identical to the process of mean-centering, if individuals are well-calibrated with regard to where they stand relative to their peers, the *average* predicted self-peer discrepancy should also be around 0. That is, individuals who are restricting their behaviors less than peers should report being lower than peers, and those who are restricting more should report being higher than peers, and this would even out in an average group estimate of 0. If this estimate were higher than 0, this would mean that, on average, individuals are overestimating what they are doing relative to their peers. We found that predicted self-peer difference in behavioral restriction (*M* = 1.75, *SD* = 1.54) was much larger than actual self-peer difference in behavior restriction (*M* = .002, *SD* =1.06), *t*(1021) = 44.51, *p* <.001, *d* = 1.39. This indicates that individuals are significantly overestimating the extent to which they are restricting their behaviors relative to their peers. However, we found that predicted behavioral restriction and actual behavioral restriction were significantly and positively correlated, *r*_*s*_ = .549, 95% CI: [.505, .590], *p* < .001, such that individuals who thought that they were restricting behavior more than their peers generally were.

Taken together, this suggests that individuals are inaccurate about the *size* of the asymmetry between self and peer (as indexed by the large average discrepancy between actual and perceived behavioral restrictions) but accurate about the *direction* of the asymmetry (as indexed by a positive relationship between predicted and actual behavioral restriction asymmetries).

#### Relationship between perceived behavioral restriction and actual behavior change

To examine whether beliefs about the extent to which individuals restricted their behavior relative to their peers related to actual differences in behavior change, we examined self-peer differences in the number of activities that participants had reported limiting over the past two weeks. We found that on average, participants reported limiting 14.75 (*SD* = 3.58) behaviors. We found that predicted behavioral restriction relative to one’s peers was positively correlated with the number of behaviors one reported limiting relative to peer group averages, *r* = .28, *t*(1020) = 9.30, *p* < .001.

### Linking self-other asymmetries in Covid-19 knowledge and behavior to mental health outcomes (Hypothesis 3)

#### Relationship between self-peer asymmetries and mental health outcomes

Next, we examined the role that perceived self-peer asymmetry may play in maladaptive emotional responding. To do so, we quantified the relationship between asymmetry measures (knowledge asymmetry, behavioral restriction asymmetry) and both Covid-specific worry, stress, and anxiety more generally.

##### Knowledge asymmetries and mental health outcomes

Turning first to Covid-specific worry, we found that, on average, our participants were moderately worried about Covid-19, *M* = 3.53, *SD* = 1.23. Consistent with our hypothesis, we found that individuals who felt that they were more knowledgeable about Covid-19 were also those that reported a higher frequency of Covid-specific worry, *b* = .21, *SE* = .03, 95% CI: [.149, .278], *t*(1021) = 6.53, *p* < .001. To examine whether this asymmetry predicted Covid-19 worry specifically or stress and anxiety more generally, we also examined the relationship between knowledge asymmetry and stress (as assessed by the PSS) and reported anxiety (as assessed by the DSM-CC-5). We did not find a relationship between knowledge asymmetry and experienced stress, *b* = -.07, *SE* = .19, 95% CI: [-.448, .305], *t*(1021) = .37, *p* = .71. We did, however, find a marginally significant relationship between Covid knowledge asymmetry and anxiety, *b* = .06, *SE* = .03, 95% CI: [-.002, .123], *t*(1021) = 1.90, *p* = .057.

##### Behavioral restriction asymmetries and mental health outcomes

We found that Covid-specific worry was significantly associated with the predicted self-peer difference in behavioral restriction, *b* = .26, *SE* = .04, 95% CI: [.189, .340], *t*(1021) = 6.90, *p* < .001. We also found a significant relationship between stress and predicted self-peer behavioral restriction, *b* = .04, *SE* = .01, 95% CI: [.024, .05], *t*(1021) = 5.49, *p* < .001, and between anxiety and predicted self-peer behavioral restriction, *b* = .25, *SE* = .04, 95% CI: [.173, .331], *t*(1021) = 6.26, *p* < .001. Taken together, this suggests that individuals who perceived greater differences in behavioral restriction between themselves and their peers are also more worried about Covid-19, and experiencing greater stress and anxiety.

##### Testing whether perceived self-peer asymmetry mediates the relationship between Covid-specific worry and general anxiety

Next, we examined whether a larger self-other asymmetry in perceived behavioral restrictions is linked to more negative mental health outcomes during the Covid-19 lockdown period. Specifically, we were interested in whether anxiety symptoms arose through the experiences of Covid-specific worry and stress during the pandemic by way of the perception that one personally took more action to both reduce likelihood of personal infection and transmission of the virus.

To do so, we tested two indirect effects models wherein self-other perceived behavioral asymmetry acted as mediators between 1) Covid-specific worry and 2) stress as they relate to more general anxiety (all model variables standardized; Figures 3 and 4). We tested these pathways as compared to the reverse (i.e., general anxiety predicting Covid-specific worry or stress) as testing reverse pathways is limited in its ability to identify feasible models (Thoemmes, 2015), and our proposed pathway is grounded in theoretical basis since prior research has indicated that the lockdown period has been associated with global increases in anxiety and depression (Salari et al., 2020; Huang & Zhao, 2020; Ahmed et al., 2020; Bäuerle et al., 2020). We posited that perceived asymmetries in behavioral restriction may be one of the (likely many) variables underlying this relationship. Individuals who are more concerned about Covid-19 may also believe that they are limiting their behaviors to a greater extent than peers. Given that Covid-19 spreads due to contact between individuals, this means that peer actions (which are out of one’s control) can have meaningful consequences for the health of the self and others, and that this may be associated with increased feelings of anxiety during the early lockdown period.

**Figure 3.**
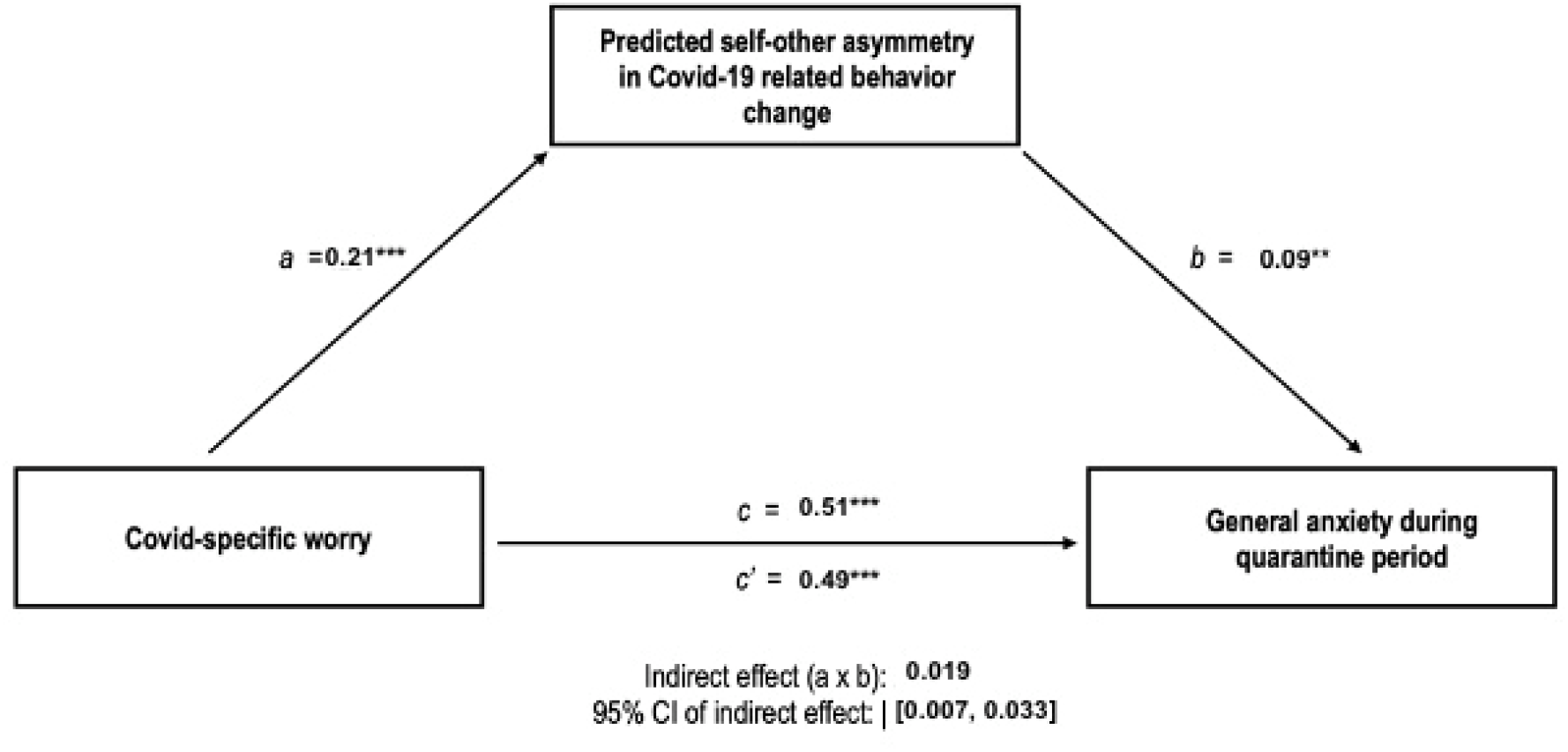
Self-other asymmetry in perceived behavioral change mediated the relationship between Covid-specific worry and general anxiety during the quarantine period.

**Figure 4.**
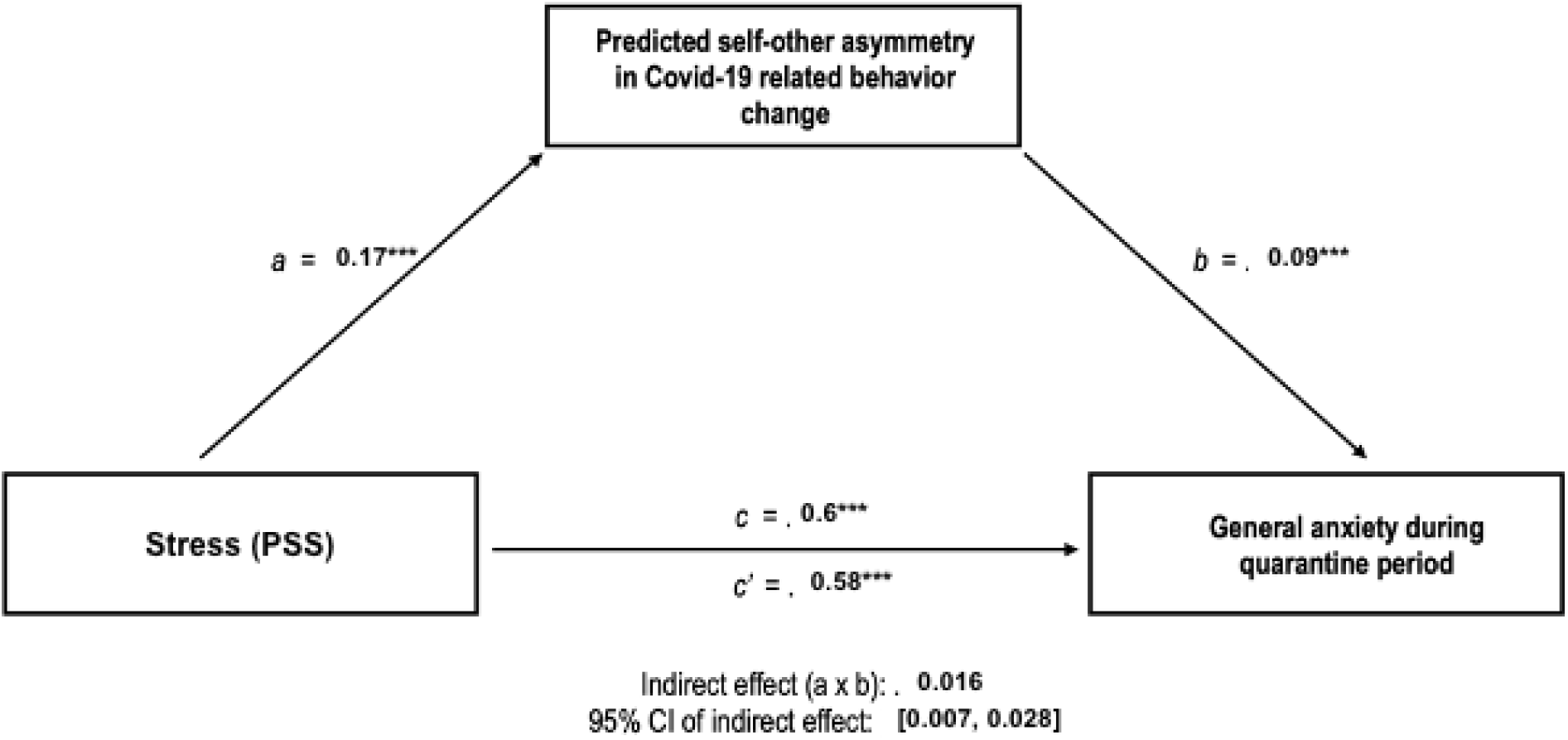
Self-other asymmetry in perceived behavioral change mediated the relationship between stress (via PSS) and general anxiety during the quarantine period.

However, we acknowledge the limitations of mediational claims/analyses—namely, that the scope of our (and any) study is limited such that we cannot capture all possible indirect effects that would impact the relationships from Covid-related worry or stress to general anxiety early in the pandemic (Fiedler, Harris, & Schott, 2018), and thus report the following in addition to the mediation analysis: 1) the correlations across all included variables so that readers may assess the association between the variables of interest (Table 2), and 2) the results of testing another potential mediator (Covid-related knowledge) assessed within the same participants at the same time. We test this additional model/alternative mediator as it is possible that individuals who are more concerned or stressed about Covid-19 may also be those who have sought out more information about it, and that having increased information about a serious disease may be associated with heightened general anxiety.

**Table 2.**
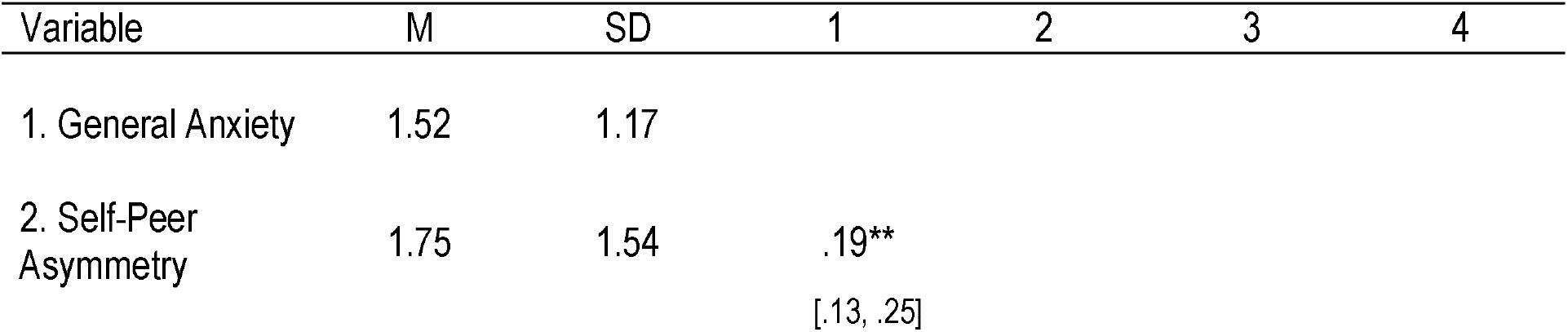
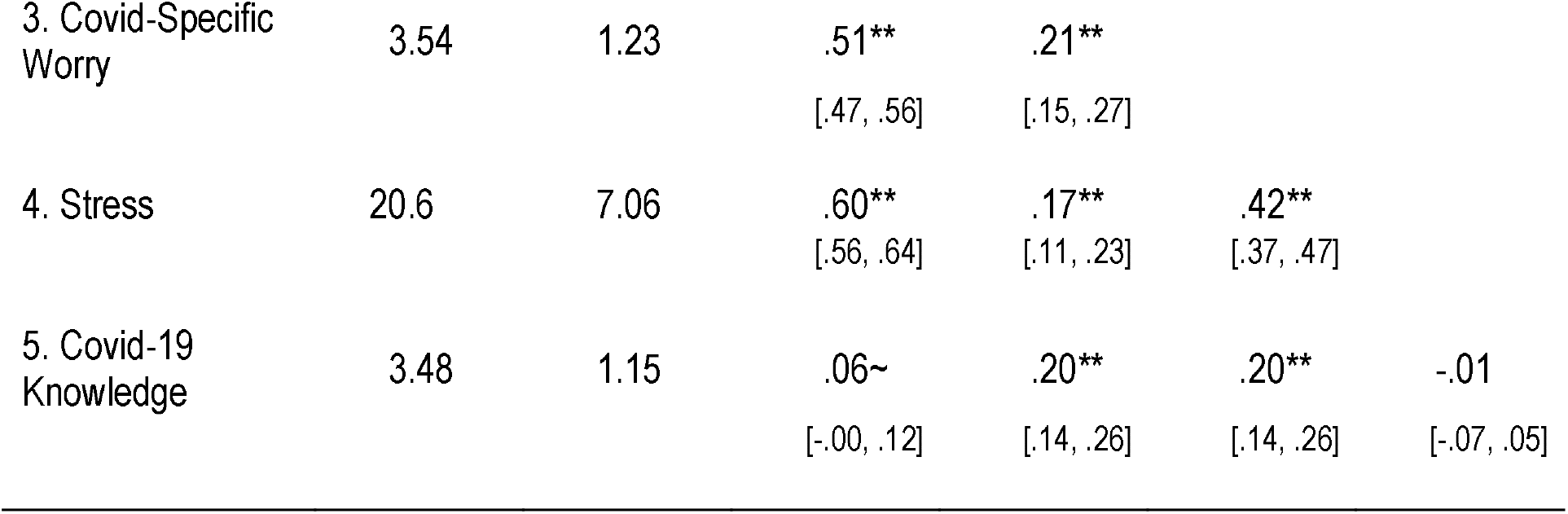
Correlations between the variables of interest. * indicates *p* < .05. ∼ indicates *p* < .06.

We found that the pairwise correlations between Covid-specific worry and self-peer perceived behavioral asymmetry (*r* = .21, *p* < .001) and between self-peer behavioral asymmetry and general anxiety (*r* = .19, *p* < .001) are strong enough to partially account for the relationship between Covid-specific worry and general anxiety (*r* = .51, *p* < .001). This is consistent with a mediation model (indirect effect determined via 5000 iterations of non-parametric bootstrapping: *b* = .019, 95% CI: [.007, .033], *SE* = 0.007, *z* = 2.79, *p* = .005), but also consistent with other causal models (Figure 3). This potentially indicates that while Covid-related worry was a strong predictor of general anxiety during the early lockdown period, this was due, in part, to the extent to which individuals felt that they were doing more than peers to reduce the spread of the virus.

Likewise, we found that the pairwise correlations between stress during the pandemic and self-peer perceived behavioral asymmetry (*r* = .17, *p* < .001) and between self-peer behavioral asymmetry and general anxiety (*r* = .19, *p* < .001) are strong enough to partially account for the relationship between stress and general anxiety (*r* = .6, *p* < .001). Again, this is consistent with a mediation model (indirect effect determined via 5000 iterations of non-parametric bootstrapping: *b* = .016, 95% CI: [.007, .029], *SE* = 0.005, *z* = 2.94, *p* = .003), but also consistent with other causal models (Figure 4). This suggests that, like anxiety, stress was also a strong predictor of general anxiety during the early stages of the Covid-19 lockdown partially by way of estimation that one took more action to reduce spread of the virus than peers.

Unlike the relationship observed for self-peer behavioral asymmetry, we did not find that the pairwise comparison between Covid-19 related knowledge and Covid-specific worry (*r* = .20, *p* < .001) and Covid-specific knowledge and general anxiety (*r* = .06, *p* = .057) was strong enough to partially account for the relationship between Covid-specific worry and general anxiety (indirect effect determined via 5000 iterations of non-parametric bootstrapping: *b* = - .009, 95% CI: [-.022, .002], *SE* = 0.006, *z* = -1.49, *p* = .137). Likewise, we found that the pairwise comparison between Covid-19 related knowledge and stress were not significantly related (*r* = -.01, *p* = 0.71), and that the pairwise comparison between those two variables and Covid-specific knowledge and general anxiety (*r* = .06, *p* = .057) was also not strong enough to partially account for the relationship between stress and general anxiety (indirect effect determined via 5000 iterations of non-parametric bootstrapping: *b* = -.001, 95% CI: [-.006, .003], *SE* = 0.002, *z* = -0.34, *p* = .732). This suggests that while differences in Covid-related knowledge is associated with Covid-specific worry and is significantly and/or marginally correlated with all of the variables of interest, it is not a promising candidate as an alternative mediator.

## Discussion

We found evidence for substantial self-peer asymmetries regarding the Covid-19 pandemic. Individuals believe (inaccurately) that they know more about Covid-19 than their peers, are less likely to contract and transmit the virus to others, are restricting their behaviors to a greater extent than their peers, and are more pro-socially motivated. We found that this self-peer asymmetry, which has been associated with positive psychological outcomes in earlier research, is instead linked to negative mental health outcomes during the Covid-19 period. The extent to which people thought they were restricting their behavior more than their peers (self-peer asymmetry) was strongly associated with both Covid-specific concern, stress, and generalized anxiety during early stages of the pandemic. Taken together, this suggests that while self-other asymmetries may be adaptive or even beneficial in typical circumstances, they may be psychologically suboptimal in collective action contexts.

We found evidence for both accuracy and inaccuracy when estimating peer knowledge and behavior. Individuals reported that they were more knowledgeable about Covid-19 relative to their peers -- a prediction not borne out in reality. Given the extent of coverage that the Covid-19 epidemic received during the data collection period, it is likely that individuals were often exposed to information about the pandemic and indeed *were* knowledgeable about the disease. However, they failed to recognize that their peers were also likely inundated by Covid-19 related news, and were likely also knowledgeable.

We also found that individuals were correct about the direction, but overestimated the magnitude of the self-peer asymmetry in behavior. Individuals believed that they were restricting their behavior more than their peers to a greater extent than was actually the case. However, individuals who were restricting their behavior more (or less) than peers were accurate about their relative standing. This suggests that individuals are correctly identifying where they stand relative to peers in terms of limiting behavior, but are perhaps too generous when it comes to determining how *far* above their peers they actually are. Consistent with accounts of unrealistic optimism (Weinstein, 1982; Dunning, Heath, & Suls, 2004), we found that individuals predicted that they were less likely than their peers to get infected with or transmit Covid-19 to others. Individuals were also more likely to claim that they were restricting their behavior for pro-social reasons than were their peers. This is consistent with research indicating that individuals tend to amplify the extent to which they are above-average in positive and moral domains (Campbell, Rudich, & Sedikides, 2002), and report being more morally motivated relative to their peers (Epley & Dunning, 2000).

While self-serving bias is often associated with maximizing happiness and self-esteem, we found that this belief pattern is associated with greater amounts of anxiety and stress in the context of Covid-19. This could be due to a number of factors. Feeling a lack of situational control is associated with increased negative affect (Thompson et al., 1993), and it is possible that participants who are more pessimistic about their peers feel less situational control in the Covid-19 context. Research has also found that while self-enhancement may facilitate coping in traumatic and negative situations (Bonanno, Field, Kovacevic, & Kaltman, 2002), it is also associated with being less liked by others (Bonanno, et al., 2002; Colvin, Block, & Funder, 1995; Joiner et al., 2003; Paulhus, 1998). It is possible that individuals who exhibited greater self-other asymmetry have less social support, increasing anxiety during a stressful time. Both situational control and perceived social support should be examined in future research as causal factors.

### Limitations

One limitation is the ambiguity about whom participants were thinking of when asked about an “average peer”. Although this framing is fairly consistent with past research on self-other asymmetries (e.g., prediction of personal behavior and that of an anonymous peer or broadly-construed “peers”, Epley & Dunning, 2000), some prior work has found that self-other asymmetries are mitigated as targets become more individuated (Alicke et al., 1995), and it is likely that these results may not be as pronounced if individuals compared themselves to an individuated peer. Although they were members of the same university community which gives way to formation of many relationships and shared experiences (e.g., exposure to the same university Covid-19 communications and regulations), we cannot be completely certain that individuals were thinking of their university-specific peers specifically when making self-peer assessments. Although university peers would most likely be a salient peer community, it is possible that participants were perhaps comparing themselves to their hometown friends, Instagram followers, or the neighbors down the street. It is worth noting that this would only change the results with regard to self-peer accuracy, and not the general conclusions with regard to mental health outcomes.

It is also unclear what information motivated the self-enhancing assessments observed here. Many factors have been identified as underlying self-enhancement, including failure to integrate base-rate information when evaluating the self (Epley & Dunning, 2000), a failure to receive or integrate objective feedback (Dunning, Heath, & Suls, 2004), not having the skills or expertise to be able to recognize one’s own incompetence (Kruger & Dunning, 1999), and motivated autobiographical recall (Helion, Helzer, Kim, & Pizarro, 2020). It is possible that when assessing peer behaviors individuals tended to rely more on statistical information about things like mask compliance, and when assessing their own behaviors instead selectively recalled times that they socially distanced or social invitations that they declined. It’s also likely that rational factors give rise to this bias — individuals have more information about the extent to which they have limited or changed behavior following the Covid-19 epidemic as compared to the information they have about their peers.

Finally, it is also worth noting the limitations of mediation analysis in terms of identifying causal mechanisms – as mentioned in our results, there are many potential mediators in the relationship between Covid-19 specific worry and anxiety (e.g., financial stress, job loss, health concerns), and identifying perceived self-other asymmetry as a potential mediator between Covid-19 specific worry and general anxiety does not preclude the existence or identification of other potential mediators.

To our knowledge, this is the first study to examine self-other asymmetry in the context of a collective action problem. To this end, it is also worth noting that our results come from a sample taken from a university located in the United States and are thus limited in their generalizability in that regard. This is particularly true given known differences about Covid-19 responses and attitudes in the U.S. and other nations (e.g., trust in others, Lenton, Boulton, & Scheffer, 2022). Future research should examine the long-term effects of self-enhancement related anxiety -- does it function as a motivator for pro-social action, or does it increase the likelihood of burnout? Given that humans will face no shortage of collective action problems in the foreseeable future (racial injustice, climate change, mass extinctions) understanding how the psychological processes that we study in the lab promote, prevent, and preclude collective action has perhaps never been so important.

## Data Availability

Data and scripts are available via OSF.

https://osf.io/hpv43/?view_only=fec42fadb67240aa8d3c8780d002bc3f

## Open Practices

Analyses -- unless indicated otherwise -- were preregistered (https://aspredicted.org/blind.php?x=/HGQ_8XB). All data and analysis scripts/output are available via Open Science Framework (OSF): https://osf.io/hpv43/?view_only=fec42fadb67240aa8d3c8780d002bc3f.

## Author contributions (CRediT)

C.H., D.V.S., and J.J. were responsible for conceptualization. J.J. oversaw data curation and project administration. C.H. and V.U. was responsible for formal analysis and visualization. C.H. was responsible for original draft writing. J.J., V.U., and D.V.S. oversaw review and editing of writing.

